# Geospatial assessment of racial/ethnic composition, social vulnerability, and lead service lines in New York City

**DOI:** 10.1101/2022.10.11.22280958

**Authors:** Anne E. Nigra, Wil Lieberman-Cribbin, Benjamín C. Bostick, Steven N. Chillrud, Daniel Carrión

**Author notes:** **Contact/Corresponding Author:** Anne Nigra, ScM, PhD, Assistant Professor, Department of Environmental Health Sciences, Columbia University Mailman School of Public Health, 722 W 168th St, 11th Floor Rm 1107, New York, NY 10032.

## Abstract

**Background:** The state of New York expects to receive $115 million in 2022 alone from the US Infrastructure Investment and Jobs Act to support the replacement of lead water service lines.

**Objectives:** To determine the number and proportion of *Potential Lead* water service lines across New York City (NYC), and the association between racial/ethnic composition, housing vulnerability, and child lead exposure vulnerability with service line type (*Potential Lead, Unknown*) at the census tract level for N= 2,083 NYC tracts.

**Methods:** We used conditional autoregressive Bayesian Poisson models to assess the relative risk (median posterior estimates, and 95% credible interval, CrI) of service line type per 20% higher proportion of residents in a given racial/ethnic group, and per higher housing vulnerability and child lead exposure vulnerability index scores corresponding to the interquartile range. We also evaluated the associations in flexible natural cubic spline models.

**Results:** Out of 854,672 residential service line records, 136,891 (16.0%) were *Potential Lead* and 227,443 (26.6%) were *Unknown*. In fully adjusted models, higher proportions of Hispanic/Latino residents and higher child lead exposure vulnerability were associated with *Potential Lead* service lines in flexible spline models and linear models (RR 1.15, 95% CrI 1.11, 1.21, and RR 1.11, 95% CrI 1.02, 1.20, respectively). Associations were modified by borough; *Potential Lead* service lines were associated with higher proportions of non-Hispanic White and non-Hispanic Asian residents in the Bronx and Manhattan, and with higher proportions of non-Hispanic Black residents in Queens.

**Discussion:** NYC has a high number of *Potential Lead* and *Unknown* service lines. Communities with a high proportion of Hispanic/Latino residents and those with children who are already highly vulnerable to lead exposures from numerous sources are disproportionately impacted by *Potential Lead* service lines. These findings can inform equitable service line replacement across NY state and NYC.

## INTRODUCTION

Ongoing racial/ethnic disparities in water lead concentrations, lead exposure from household dust and paint, and child blood lead levels (a measure of internal dose) in the US are well documented and are of great public health concern.^1-4^ Substantial epidemiologic and toxicologic evidence supports that there is no safe level of lead exposure, especially for children.^5-8^ Large scale infrastructural investments are needed to further reduce disparities and to decrease overall exposure to historical sources of lead, many of which are still present in the built environment.

Lead enters public drinking water via corrosion of distribution and plumbing system materials that contain lead, including solder, fixtures, faucets, and lead service lines that connect homes to public drinking water distribution systems (acidity, mineral content, and other source water characteristics also influence corrosivity).^9^ Although the US EPA Lead and Copper Rule (LCR) requires public water systems to conduct routine compliance monitoring for lead at taps throughout the distribution system and to intervene to reduce lead corrosion, current regulations are inadequate to reduce water lead exposure and eliminate racial/ethnic disparities in water lead exposure. For example, the current LCR fails to establish a health-based standard, enables the biased selection of taps and homes selected for sampling (resulting in documented racial disparities in reported compliance monitoring sample measurements), and does not require the reporting of absolute measured concentrations at all monitoring points (only higher percentile values are reported when measured above the action level) even though water lead concentrations are highly variable across season and within water distribution systems.^1,10-14^ Large scale replacement of lead water service lines, which are significant sources of lead to drinking water even when source water contains no lead, remains a critically needed intervention to reduce public drinking water lead exposures and disparities.^9^

In the fall of 2021, the US Congress passed the Infrastructure Investment and Jobs Act, which allocates $55 billion to improving drinking water quality across the US.^15^ New York state (NYS) expects to receive $115 million in 2022 alone to support the replacement of lead water service lines.^16^ Identifying communities with higher prevalence of lead service lines is critically needed to help inform equitable prioritization of lead service line replacement throughout New York City (NYC) and NYS. Under NYS’s current lead service line replacement program, municipalities are prioritized for funding if those municipalities are low income (median household income <150% of regional average), are likely to have a high number of lead services lines (500 or more homes constructed before 1939), and have a high percentage of children with elevated blood lead levels (>0.5% children with blood level >5 μg/dL, although the Centers for Disease Control and Prevention’s (CDC) level is 3.5 μg/dL).^17^ Although the location of lead service lines is largely unknown for most utilities, the NYC Department of Environmental Protection (NYCDEP) recently published a database of water service line material for city buildings (water service line material is recorded as *Potential Lead, Unknown, Not Lead*, and *Not Applicable*).^18^ The database provides a unique and powerful opportunity to evaluate neighborhood level sociodemographic characteristics with potential lead service lines. Integrating this environmental information with community racial/ethnic composition and other domains of social vulnerability are needed to prioritize activities within NYC and other large municipalities.

Our objectives were to a) to characterize the number and percentage of *Potential Lead* and *Unknown* service lines in NYC overall and across political jurisdictions, and b) to evaluate the association between racial/ethnic composition and reported water service line material (specifically *Potential Lead* and *Unknown* service lines, as we are specifically interested in lead line replacement) across NYC at the census tract level using geospatial models. In secondary analyses, we also evaluated the association between two constructed measures of social vulnerability (housing vulnerability and child lead exposure vulnerability) and service line material. In this paper, we evaluate neighborhood racial/ethnic composition to inform equitable interventions and consider that differences in service line material by racial/ethnic composition reflects structural racism. Structural racism “encompasses racist practices and systems that are imbedded into institutions in the US,” and is the “totality of ways in which societies foster racial discrimination through mutually reinforcing systems.^19,20^” The leading framework explaining how racial/ethnic and other sociodemographic disparities in US drinking water exposures are created and reinforced theorizes that inequities in the natural (e.g. hydrogeology, climate), built (e.g. infrastructure, technical and financial capacity, economies of scale), and sociopolitical (e.g. social and political capital, housing discrimination) environments underlie these disparities.^21^ For drinking water lead exposure, inequities in water infrastructure, housing discrimination, and inadequacies in federal drinking water regulations are particularly relevant.

## METHODS

### New York City lead service lines and building-level data

NYCDEP’s Lead Service Line Location Coordinates database^18^ was published in 2019 and contains a water service line record (coded as *Potential Lead, Unknown, Not Lead*, or *Not Applicable)* for 857,536 buildings across NYC (>99.9% of tax lots in NYC). Service line types are largely determined by historical records (e.g. latest plumbing record filed by a licensed plumber), and are supplemented by observational and external excavation records (e.g. inspections inside the home, observations of the connection to the main line). NYCDEP explicitly does not guarantee the accuracy or completeness of the database, which is updated semi-annually.^18^

To identify buildings with rental units, we merged the water service line records with the NYC Department of Housing and Preservation’s Multiple Dwelling Unit Registry (these include residential buildings with three or more units, and one- or two-family homes that are not owner-occupied).^22^ We also merged in the NYC Primary Land Use Tax Lot Output (PLUTO) dataset, which contains information on building build year and geographic location.^23^ We aggregated building-level data to the census tract level and calculated the total number and proportion of water service lines recorded as *Potential Lead* and *Unknown* per tract. We aggregated to the census tract because the main sociodemographic variables of interest were only available at the census tract or county level, and the primary purpose of our analysis was to identify characteristics of communities (not buildings) with a higher proportion of *Potential Lead* service lines.

### Census tract sociodemographic characteristics

We downloaded census tract level sociodemographic characteristics from the 2018 US Census Bureau’s American Community Survey (five-year estimates) via the *tidycensus* package in R, including: total population and number of households; median household income and rent; number of households receiving supplemental nutrition assistance program (SNAP) benefits; number of residents aged 25-64 without a high school diploma; number of adult residents without health insurance; number of residents living below 150% of the federal poverty line; number of overall, vacant, occupied, owner occupied, and renter occupied housing units; number of unemployed residents, and the proportion of residents who were non-Hispanic Black, non-Hispanic White, Hispanic/Latino, non-Hispanic Asian, and non-Hispanic American Indian.^24^

### Housing vulnerability and child lead exposure vulnerability indices

We developed NYC-specific indices of housing vulnerability and child lead exposure vulnerability for secondary analyses. For both indices, each census tract was ranked for each of the individual variables comprising the index, and these ranks were summed to generate the final index score on a continuous scale. This approach matches widely used indices of social vulnerability, including the Center for Disease Control and Prevention (CDC)’s nationwide social vulnerability index.^25^ Our housing vulnerability index was specifically designed to characterize housing insecurity and/or instability, and was derived from the following variables: housing burden (median income divided by median rent), the percent of occupied residential units that were rented, and the proportion of residents receiving SNAP benefits, living below 150% of the federal poverty line, and 25-64 years of age without a high school diploma.

Our child lead exposure vulnerability index was developed specifically to characterize NYC areas where children could be highly vulnerable to lead exposure from all potential sources (e.g. paint, water, dust), and was derived from the following variables: the number of residents <6 years old, median household income, proportion of residential units that were vacant (proxy for dilapidation/disrepair), the proportion of occupied residential units that were rented, and the proportion of residents receiving SNAP benefits, without health insurance, and 25-64 years of age without a high school diploma. We did not include median building construction year in either index to separately assess the impact of further adjusting for housing age (relevant for the presence of lead paint and dust) in progressively adjusted models. Because these indices contain several common variables and are highly correlated (Pearson’s rho=0.94, p-value<0.001), differences in findings across these two vulnerability indices are likely explained by non-overlapping variables (i.e. the number of residents <6 years old, median household income, the proportion of residential units that were vacant, and the proportion of adults without health insurance) or unmeasured/unanalyzed variables that correlate with them.

To conduct sensitivity analyses using previously developed indices of social and socioeconomic vulnerability, we downloaded the nationwide community deprivation index published by Brokamp et. al and the CDC’s nationwide social vulnerability sub-index for socioeconomic vulnerability.^25,26^ We did not assess the CDC’s overall social vulnerability index because the Housing Type and Transportation sub-index is not relevant for NYC and the Language and Minority Status sub-index would over adjust for racial/ethnic composition.

### Exclusion criteria

Out of 857,536 residential building service line records available in the NYC Lead Service Line Location Coordinates dataset, we excluded 2,864 records (0.3%) which could not be matched the PLUTO dataset by the building’s Borough, Block, and Lot number for a final N of 854,672 buildings. Out of 2,165 NYC census tracts, we excluded 42 with a population of 0 (e.g. parks, cemeteries), 35 missing the housing vulnerability and child lead exposure vulnerability index score (22 were missing median household income and 13 were missing housing burden), and 5 missing median building construction year for a total analytic sample of N= 2,083 census tracts. No census tracts were missing water service line material or racial/ethnic composition variables.

### Descriptive statistical analysis

All data management and statistical analysis was conducted in R version 4.1.1. To characterize the burden of *Potential Lead* and *Unknown* service lines in NYC overall and across political jurisdictions, we tabulated the total number and proportion of water service lines by material *(Potential Lead, Unknown, Not Lead*, and *Not Applicable)* stratified by various political jurisdictions (zip code, community district, borough, state senate district, and state assembly district) and further stratified by building characteristics (city-owned buildings and multiple dwelling buildings). At the census tract level, we first evaluated the distribution of service line records, population density, racial/ethnic composition, and sociodemographic characteristics including index scores of housing vulnerability and child lead exposure vulnerability for all census tracts within NYC and across quartiles of the proportion of water service lines recorded as *Potential Lead*.

### Conditional autoregressive Bayesian Poisson regression

We evaluated spatial autocorrelation in the rates of *Unknown* and *Potential Lead* water service lines across census tracts by visually assessing the residuals from Poisson regression models and by assessing Moran’s I (correlation coefficient measuring spatial autocorrelation). Spatial neighbors were identified via a queen contiguity matrix for neighbors (*i*=1) versus non-neighbors (*i*=0). Moran’s I indicated positive global spatial autocorrelation (*Potential Lead:* I = 0.58, p<0.001; *Unknown:* I= 0.52, p<0.001; local indicators of spatial association (LISA) clusters are mapped in **Supplemental Figure 1**). We therefore implemented globally smooth, conditional autoregressive Bayesian Poisson regression via the *S.CARbym* function in the ‘CARBayes’ R package, with census tract as the unit of analysis.^27^ For all models, the offset term was set to the natural log of the number of buildings per census tract with service line material record available. For all analyses, we focused on *Potential Lead* and *Unknown* water service lines (rather than *Not Lead* lines) because we are specifically interested in community characteristics for prioritization of lead service line replacement. Model results and maps of spatial residuals showed no evidence of residual spatial autocorrelation for our main analyses (**Supplemental Figure 2**).

To determine if racial/ethnic composition was associated with service line type, we assessed the relative risk (RR, posterior median estimates) and corresponding 95% credible intervals (CrIs) of *Potential Lead* and *Unknown* service lines per 20% higher proportion of residents in each racial/ethnic subgroup (absolute change, e.g. from 20% to 40%). We were unable to assess associations for American Indian/Alaskan Native residents because the census tract level proportion of residents in this racial/ethnic group was very small and often zero (mean 0.2%). Because the interquartile range of census tract level percentage of residents in a given racial/ethnic group was 36% for non-Hispanic Black, 54% for non-Hispanic White, 30% for Hispanic/Latino, and 18% for non-Hispanic Asian residents, we chose 20% as our primary unit of change to enable direct comparison across models assessing higher proportions of residents in different racial/ethnic groups. Model 1 adjusted for population density and the child lead exposure vulnerability index. Model 2 further adjusted for median building construction year to assess the impact of further adjustment for housing age (to evaluate whether the association between racial/ethnic composition and service line material is explained by differences in the build year of neighborhood housing stock). To account for potential nonlinearity in the association, we also evaluated the association in flexible natural cubic spline models with knots at the 33^rd^ and 66^th^ percentiles and the reference set to the 10^th^ percentile of the distribution. We also stratified our main linear analyses by borough (Bronx, Brooklyn, Manhattan, Queens, Staten Island) because racial/ethnic composition differs substantially across and within NYC boroughs.

To determine if our NYC-specific indices of housing vulnerability and child lead exposure vulnerability were associated with service line type (secondary analyses), we assessed the RR and corresponding 95% CrIs of *Potential Lead* and *Unknown* service lines per an increase in each index score corresponding to the interquartile range. Model 1 adjusted for population density and racial/ethnic composition. Model 2 further adjusted for median building construction year, also to assess the impact of further adjustment for housing age. We also repeated these analyses with flexible spline models to assess potential non-linearity.

### Sensitivity analyses

We performed several additional sensitivity analyses. First, we modeled the RR of service line type per a 10% higher proportion of residents in each racial/ethnic subgroup (instead of 20%), with similar findings but slightly attenuated effect estimates (**Supplemental Table 1**). For models evaluating racial/ethnic composition, we replaced adjustment for the childhood lead exposure vulnerability index with adjustment for three different indices of social vulnerability and socioeconomic status, including the NYC-specific housing vulnerability index we developed, the CDC’s index for socioeconomic vulnerability, and the community deprivation index developed by Brokamp et. al, all with similar findings (**Supplemental Table 2**). To determine if census tracts with very few service line records influenced our findings, we repeated our main analyses after restricting to census tracts with at least 25 service line records, also with similar findings (**Supplemental Table 3**). We also assessed the RR and corresponding 95% CrIs of *Potential Lead* and *Unknown* service lines across quartiles of each social vulnerability index, comparing the second, third, and fourth quartile to the first (reference) (**Supplemental Table 4)**. Finally, we used a “leave-one-out” modeling approach to evaluate if observed significant associations for higher proportions of residents of a given racial/ethnic group was related to lower proportions of residents of another specific racial/ethnic group. This modeling approach is commonly used in the nutrition and arsenic methylation literature when proportions which sum to 100% are modeled. By adjusting for the proportion of residents in all other racial/ethnic groups but leaving one group out of the model, the interpretation of the effect estimate is per higher proportion of residents in the group of interest specifically because the proportion of residents in the group left out was lower.^28,29^

## RESULTS

Out of 854,672 service lines, 489,535 (57.3%) were reported as *Not Lead*, 136,891 (16.0%) as *Potential Lead*, 227,443 (26.6%) as *Unknown*, and 803 (<1%) as *Not Applicable* (**Supplemental Table 5**). The largest absolute number of *Potential Lead* and *Unknown* service lines were in Queens (N= 66,508) and Brooklyn (N= 91,377), respectively (**Supplemental Table 6**). Relative to other boroughs, a larger proportion of service lines were *Potential Lead* in the Bronx (21.2%) and Queens (20.5%), and Brooklyn had the largest proportion of service lines that were *Unknown* (33.2%). Buildings with multiple dwelling units had a similar proportion of *Potential Lead* and *Unknown* services lines compared to other buildings (17.7% versus 15.6%, and 25.4% versus 26.9%, respectively) (**Supplemental Table 7**), while buildings owned by NYC had a higher proportion of *Unknown* service lines compared to other buildings (62.2% versus 26.0%) (**Supplemental Table 8**). Tables of the absolute number and proportion of service line types by zip code, community district, state senate district, and state assembly district overall and further stratified by buildings with multiple dwelling units or those which are owned by NYC are in **Supplemental Tables 9-22**.

**Figure 1** displays census tract proportions of *Unknown* and *Potential Lead* service lines. Census tracts with the highest proportion of *Unknown* service lines were located in Staten Island and southern Queens, while the highest proportion of *Potential Lead* service lines were located in eastern Queens and eastern Bronx (these areas generally have higher proportions of non-Hispanic Asian and Hispanic/Latino/non-Hispanic Black residents, respectively, see **Supplemental Figure 3**). The mean number of service line records within census tracts was 409 (minimum N=1, maximum N= 3,252), and the proportion of *Potential Lead* service lines ranged from 0% (N= 140 tracts) to 70% (N= 1 tract) (**Table 1**). Census tracts in the highest quartile of the proportion of *Potential Lead* service lines (range >23% - 70%) had lower mean total number and proportions of non-Hispanic Black and non-Hispanic White residents, higher mean total number and proportions of Hispanic/Latino residents, and higher scores of the CDC’s socioeconomic vulnerability index (**Table 1**).

**Table 1.** Characteristics of New York City census tracts overall and stratified by quartile of the proportion of water service lines recorded as *Potential Lead* (N= 2,083).

|  | Overall<br>0% - 70% | Quartile 1<br><8% | Quartile 2<br>8% - 14% | Quartile 3<br>>14% - 23% | Quartile 4<br>>23% - 70% |
| --- | --- | --- | --- | --- | --- |
| N census tracts | 2,083 | 521 | 538 | 528 | 496 |
| Mean N water service line records (min, max) | 409 (1, 3252) | 485 (1, 3252) | 351 (7, 2195) | 378 (6, 1681) | 425 (4, 1534) |
| Population density<sup>a</sup> (mean, SD) | 20,232 (13766) | 17,284 (13,917) | 22,499 (14,632) | 21,516 (13,371) | 19,504 (12,406) |
| *Racial/ethnic composition (mean N, mean %)* |  |  |  |  |  |
| non-Hispanic Black | 88,092 (23.3%) | 111,251 (29.0%) | 90,389 (21.6%) | 74,622 (19.7%) | 75,613 (22.8%) |
| non-Hispanic White | 129,082 (32.1%) | 150,201 (34.6%) | 140,866 (33.9%) | 142,485 (35.5%) | 79,848 (24.1%) |
| Hispanic/Latino | 117,344 (27.2%) | 86,143 (19.5%) | 129,101 (27.9%) | 116,772 (27.6%) | 137,972 (33.8%) |
| non-Hispanic Asian | 55,728 (14.3%) | 62,481 (14.4%) | 53,336 (14.1%) | 56,221 (14.1%) | 50,705 (14.4%) |
| non-Hispanic American Indian | 707 (0.2%) | 791 (0.2%) | 574 (0.1%) | 700 (0.2%) | 769 (0.3%) |
| *Socioeconomic characteristics (mean, SD)* |  |  |  |  |  |
| Median household income (\$) | 67,078 (33,027) | 69,614 (36,196) | 65,530 (35,916) | 68,859 (34,328) | 64,195 (23,163) |
| Average household size | 2.86 (0.66) | 2.75 (0.68) | 2.82 (0.65) | 2.78 (0.58) | 3.10 (0.66) |
| Housing burden<sup>b</sup> | 0.03 (0.01) | 0.02 (0.01) | 0.03 (0.01) | 0.03 (0.01) | 0.03 (0.01) |
| CDC socioeconomic vulnerability index<sup>c</sup> | 0.60 (0.27) | 0.55 (0.28) | 0.63 (0.29) | 0.58 (0.28) | 0.64 (0.24) |
| Community deprivation index<sup>d</sup> | 0.39 (0.14) | 0.37 (0.15) | 0.41 (0.15) | 0.39 (0.14) | 0.40 (0.12) |
| NYC housing vulnerability index<sup>e</sup> | 2.41 (1.19) | 2.14 (1.22) | 2.63 (1.24) | 2.44 (1.16) | 2.44 (1.09) |
| NYC child lead exposure vulnerability index<sup>f</sup> | 2.89 (1.10) | 2.56 (1.15) | 3.08 (1.09) | 2.94 (1.09) | 3.00 (1.00) |

<sup>a</sup> Persons per square kilometer. <sup>b</sup> Median rent divided by median income. <sup>c</sup> CDC/ATSDR socioeconomic status index includes measures of below poverty, unemployed, income, and no high school diploma. <sup>d</sup> Community deprivation index includes measures of median income, fraction vacant housing, fraction no health insurance, fraction high school education, and fraction with assisted income (Brokamp et al. 2019). <sup>e</sup> Developed for this analysis, includes NYC-wide ranking of housing burden, proportion of residents receiving supplemental nutritional assistance program (SNAP) benefits, proportion of residents below 150% of the federal poverty level, proportion of residents with no high school diploma, and proportion of occupied residential units that are rented. <sup>f</sup> Developed for this analysis, includes NYC-wide ranking of proportion of residents receiving SNAP benefits, proportion of residents with no high school diploma, median income, proportion of residents without health insurance, percent of residential housing units that are vacant, percent of occupied residential units that are rented, and total number of residents under the age of 6.

**Figure 1.**
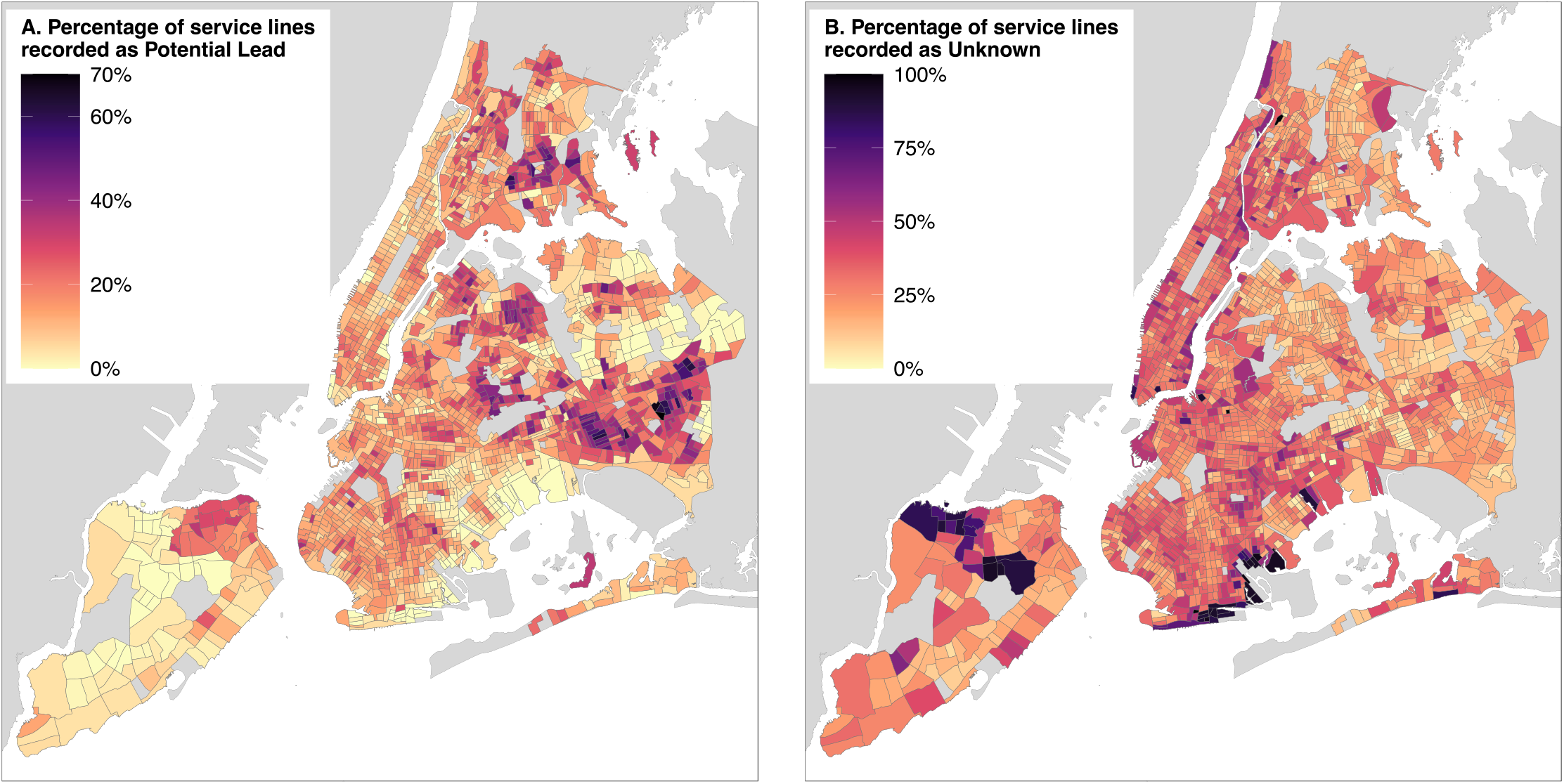
Percentage of building water service lines recorded as *Potential Lead* (Panel A) and *Unknown* (Panel B) by census tract across New York City (N= 2,083). Census tracts shown in gray were not included in the analysis. For *Potential* Lead service lines, local indicators of spatial association (LISA) clusters categorized as “High/High” were identified in northern and eastern Queens and eastern Bronx (**Supplemental Figure 1**).

**Table 2** presents the RR (95% CrIs) of *Potential Lead* and *Unknown* service lines per 20% higher proportion of residents belonging to a given racial/ethnic group at the census tract level (N= 2,083). In fully adjusted models (Model 2), the RR (95% CrI) of *Potential Lead* service lines was 0.92 (0.89, 0.96) for non-Hispanic White residents, 1.15 (1.11, 1.21) for Hispanic/Latino residents, and null for non-Hispanic Black and non-Hispanic Asian residents. For *Unknown* service lines, the RR (95% CrI) was 1.03 (1.01, 1.06) for non-Hispanic Black residents, 1.04 (1.01, 1.07) for non-Hispanic White residents, 0.94 (0.91, 0.97) for non-Hispanic Asian residents, and 0.95 (0.92, 0.98) for Hispanic/Latino residents. Flexible cubic spline models indicated that most associations were non-linear, especially for *Unknown* lines and *Potential Lead* lines for Hispanic/Latino and non-Hispanic Asian residents (**Figure 2**). In “leave-one-out” models, associations for non-Hispanic Black and non-Hispanic White residents and *Unknown* lines were driven by lower proportions of Hispanic/Latino residents (GMR 1.07, 95% CI 1.03, 1.11, and GMR 1.08, 95% CI 1.04, 1.12, respectively); associations for Hispanic/Latino and non-Hispanic Asian residents were driven by lower proportions of non-Hispanic White residents (GMR 1.16, 95% CI 1.10, 1.25 and GMR 1.08, 95% CI 1.02, 1.14, respectively).

**Table 2.** Relative risks (RR) and 95% credible intervals (CrIs) of service line type associated with racial/ethnic composition or vulnerability indices within New York City (NYC) at the census tract level (N=2,083). Models are globally smooth conditional autoregressive (CAR) Poisson models via the *S.CARbym* function in the “CARBayes” R package. Model offset is set as the log census tract total number of buildings with service line material record available. The total number of burn-in iterations was set to 20,000, 5,000 iterations were retained, and a thinning factor of 5 was applied. Spatial neighbors were identified via a queen contiguity matrix for neighbors (*i*=1) versus non-neighbors (*i*=0). We found no evidence of global spatial autocorrelation in model residuals (p>0.5, see **Supplemental Figure 2**). ^a^ Per 20% higher proportion of residents in a given racial/ethnic group. Model 1 adjusts for population density and child lead exposure vulnerability index (created from within-NYC ranking of proportion of residents receiving SNAP benefits, proportion of residents with no high school diploma, median income, proportion of residents without health insurance, percent of residential housing units that are vacant, percent of occupied residential units that are rented, and total number of children under 6 years of age). Model 2 further adjusts for median building construction year. ^b^ Per increase in index score corresponding to the interquartile range (2.0 for housing vulnerability and 1.0 for child lead exposure vulnerability). Model 1 adjusts for population density and racial/ethnic composition (proportion of residents who are non-Hispanic Black, non-Hispanic White, Hispanic/Latino, and non-Hispanic Asian). Model 2 further adjusts for median building construction year.

|  |  | *Potential Lead* | *Unknown* |
| --- | --- | --- | --- |
| *Racial/ethnic composition<sup>a</sup>* |  |  |  |
| Non-Hispanic Black | *Model 1* | 0.96 (0.92, 1.01) | 1.03 (1.01, 1.05) |
|  | *Model 2* | 0.97 (0.94, 1.01) | 1.03 (1.01, 1.06) |
| Non-Hispanic White | *Model 1* | 0.97 (0.92, 1.00) | 1.04 (1.02, 1.06) |
|  | *Model 2* | 0.92 (0.89, 0.96) | 1.04 (1.01, 1.07) |
| Hispanic/Latino | *Model 1* | 1.14 (1.09, 1.19) | 0.95 (0.91, 0.98) |
|  | *Model 2* | 1.15 (1.11, 1.21) | 0.95 (0.92, 0.98) |
| Non-Hispanic Asian | *Model 1* | 1.00 (0.95, 1.06) | 0.94 (0.91, 0.97) |
|  | *Model 2* | 1.03 (0.96, 1.08) | 0.93 (0.90, 0.95) |
| *NYC vulnerability indices<sup>b</sup>* |  |  |  |
| Housing vulnerability index | *Model 1* | 1.02 (0.82, 1.15) | 1.23 (1.15, 1.30) |
|  | *Model 2* | 1.01 (0.91, 1.09) | 1.23 (1.15, 1.31) |
| Child lead exposure vulnerability index | *Model 1* | 1.32 (1.23, 1.44) | 1.09 (1.05, 1.14) |
|  | *Model 2* | 1.11 (1.02, 1.20) | 1.09 (1.05, 1.13) |

**Figure 2.**
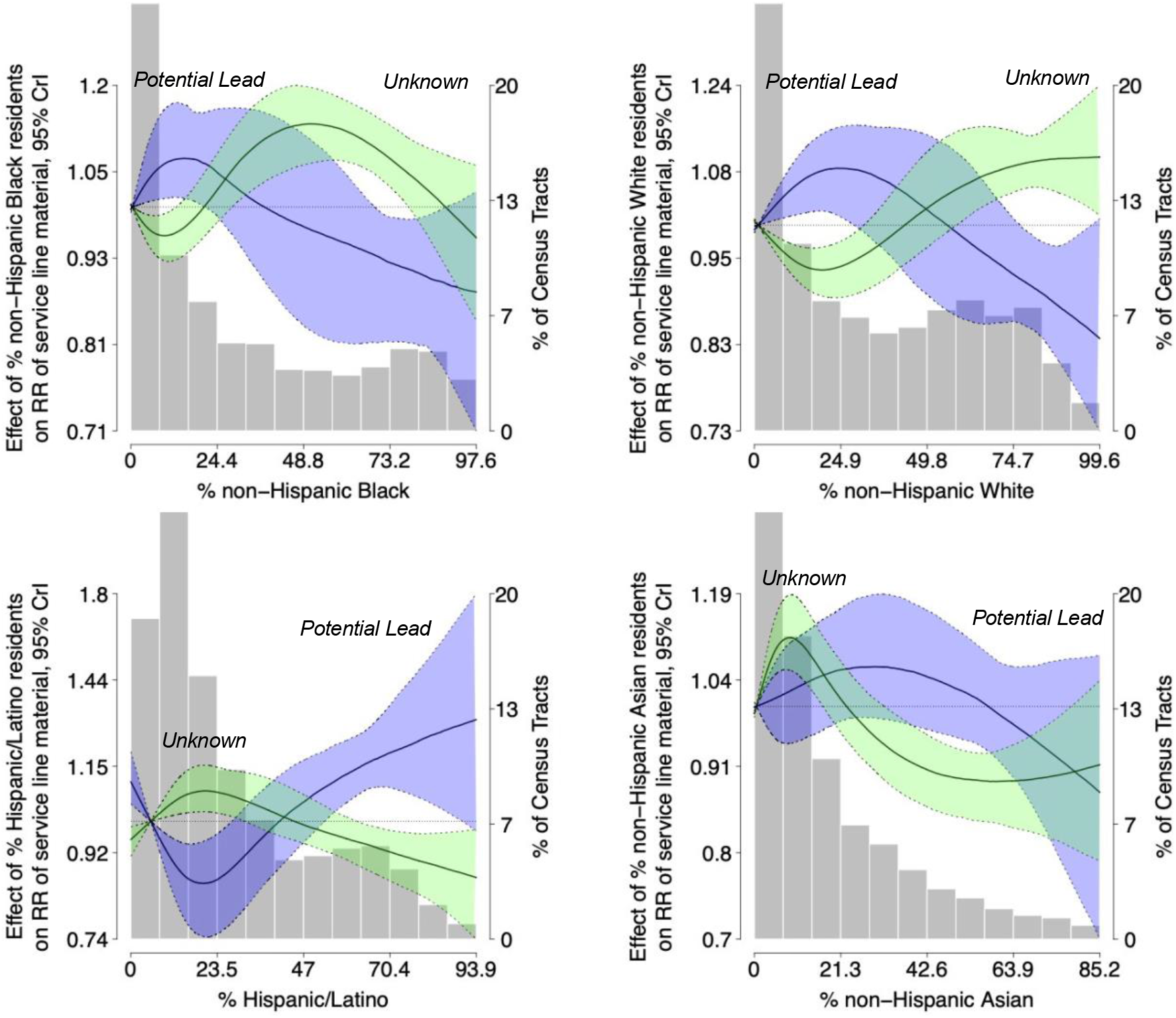
Flexible natural cubic spline models for relative risks (RR) and 95% credible intervals (CrIs) of service line type (*Potential Lead* in purple and *Unknown* in green) associated with a 20% higher proportion of residents in a given racial/ethnic subgroup within New York City at the census tract level (N=2,083). Lines represent the RR of *Potential Lead* and *Unknown* service lines by proportion of residents in a given racial/ethnic group (effect estimate values are given on the left y-axis). Green and blue shaded areas (outlined by dotted lines) represent 95% CrIs. Shaded gray bars represent the distribution of the proportion of residents in a given racial/ethnic group at the census tract level across NYC (values are given on the x-axis and the percent of census tracts with a given proportion of residents of each racial/ethnic group is given on the right y-axis). Models are globally smooth conditional autoregressive (CAR) Poisson models via the *S.CARbym* function in the “CARBayes” R package, and include knots at the 33^rd^ and 66^th^ percentiles, with the reference set to the 10^th^ percentile of the distribution. Model offset is set as the log census tract total number of buildings with service line material record available. The total number of burn-in iterations was set to 20,000, 5,000 iterations were retained, and a thinning factor of 5 was applied. Spatial neighbors were identified via a queen contiguity matrix for neighbors (i=1) versus non-neighbors (i=0). Models are adjusted for population density, median building construction year, and the constructed child lead exposure vulnerability index.

Per increase in vulnerability score corresponding to the interquartile range, higher child lead exposure vulnerability score was associated with both *Potential Lead* and *Unknown* service lines, while higher housing vulnerability score was associated with *Unknown* service lines only (**Table 2;** similar findings across quartiles, see **Supplemental Table 4**). In analyses stratified by borough, *Potential Lead* service lines were associated with higher proportions of non-Hispanic White and non-Hispanic Asian residents in the Bronx and Manhattan, and with higher proportions of non-Hispanic Black residents in Queens (**Supplemental Table 23**). For models evaluating racial/ethnic composition and vulnerability indices, further adjustment for median building construction year attenuated effect estimates for *Potential Lead* but not *Unknown* lines, reflecting that *Unknown* lines are more evenly distributed throughout the city (**Figure 1**).

## DISCUSSION

We found a high number and proportion of *Potential Lead* (16%) and *Unknown* (26.6%) service lines in residential buildings throughout NYC and significant inequities in *Potential Lead* service line prevalence, underscoring the high overall presence of lead service lines within NYC and the importance of prioritizing lead service line replacement for the most impacted communities. Higher proportions of Hispanic/Latino residents and higher child lead exposure vulnerability index scores were significantly associated with *Potential Lead* service lines. These findings suggest that NYC communities with children who are already highly vulnerable to lead exposures from numerous sources (paint, dust) have, in addition, a higher proportion of lead service lines in their communities. The positive association between child lead exposure vulnerability and *Potential Lead* service lines is likely related to variables that were not included in the housing vulnerability index (the number of residents <6 years old, median household income, the proportion of residential units that were vacant, and the proportion of adults without health insurance), or other unmeasured and correlated variables. While positive associations for Hispanic/Latino residents and non-Hispanic Asian residents with *Potential Lead* service lines was driven by lower proportions of non-Hispanic White residents, associations between non-Hispanic Black and non-Hispanic White residents and *Unknown* service lines were driven by lower proportions of *Hispanic/Latino* residents. Prioritization for service line replacement should also consider neighborhoods with higher proportions of Hispanic/Latino residents and neighborhoods with a higher proportion and absolute number of *Potential Lead* lines, including but not limited to eastern Queens and eastern Bronx, and NYC overall. An undetermined proportion of *Unknown* services lines are lead; further evaluation of these service lines is warranted throughout NYC.

These findings are supported by a growing body of literature characterizing inequities in public water system contaminant exposures across racial/ethnic and socioeconomic subgroups in the US, including lead.^4,30,31^ Inequities in public drinking water contaminant exposures result from complex, multilevel interactions between the built (water infrastructure, land use patterns), sociopolitical (social vulnerability, regulatory policies and enforcement), and natural (geologic, hydrologic conditions) environments.^21^ Racism and classism operating through the built (priorities for repairing public water infrastructure, housing age and plumbing components) and sociopolitical environments (housing discrimination, lead corrosion control policies, federal LCR regulatory policies and enforcement) are likely avenues creating and reinforcing inequities in water lead exposure. Future analyses should consider a more intersectional approach by evaluating socioeconomic vulnerability, racial/ethnic composition, and other interacting dimensions of marginalization at the neighborhood-level. Future work should also evaluate how changes in neighborhood demographic composition has changed over time in relation to the replacement of lead service lines, including assessing residential segregation.

Our findings complement known geographic, socioeconomic, and racial/ethnic inequalities in NYC child blood lead levels (which integrate all lead exposure sources including drinking water, dust, and paint) published by the NYC Department of Health and Mental Hygiene.^32-34^ For example, in 2017 the highest percentage of children with blood lead levels ≥5 μg/dL lived in Brooklyn (44%) and Queens (28%).^32^ To our knowledge, the NYC Department of Health and Mental Hygiene only makes aggregate child blood lead data publicly available at the United Healthcare Fund neighborhood level (N= 42 neighborhoods across the entire city).^35^ Deidentified, individual-level records linking child blood lead levels to housing characteristics and service line material would enable valuable analyses quantifying the absolute and relative contribution of lead sources (e.g. drinking water, diet, dust, and paint) across racial/ethnic groups in NYC.^36,37^

There are several limitations to our analyses. We relied on census tracts as our geographic unit of aggregation because the sociodemographic variables of interest for this analysis were mostly available only at the census tract and county level. Although reliance on administrative boundaries in geospatial studies can bias findings, our approach is directed at identifying community characteristics based on administrative boundaries specifically because these boundaries are likely the most useful for public health infrastructure investments and the prioritization of service line replacement, even though residents may self-define neighborhoods differently. We were unable to repeat our analysis at alternative geographic resolutions (e.g. census block, zip code) and therefore findings may differ across analyses repeated at alternative administrative units. We also relied on Census bridged race/ethnicity categories, which do not capture several important domains of race/ethnicity and limited our analysis to four crude groupings.

Our analysis is likely subject to non-trivial outcome misclassification because the NYCDEP Lead Service Line Location Coordinates dataset has not been validated and is based largely on historical records updated semi-annually. Outcome misclassification may be differential by neighborhood sociodemographic characteristics (e.g. single-family home owners in high-income neighborhoods may be more likely to contract with licensed plumbers, resulting in more accurate records). While non-differential exposure misclassification can bias findings in either direction, we expect that lead service lines were more likely replaced in buildings and neighborhoods with a higher proportion of non-Hispanic White residents, and that these records are more likely to be correctly updated in NYCDEP’s Lead Service Line Location Coordinates database. We suspect that observed associations for *Potential Lead* service lines might be underestimated for Hispanic/Latino, non-Hispanic Black, and non-Hispanic Asian groups because a higher proportion of *Unknown* lines may be lead. Future studies should assess the validity of reported service line material in a targeted, representative sample of NYC buildings.

There is no safe level of lead exposure for children.^5,38,39^ Even at lower levels of exposure, lead is associated with impaired cognitive function, attention-related behavioral problems, and diminished academic performance, with many studies indicating that the steepest declines in neurocognitive outcome measures occur at the lowest levels of blood lead.^6,40,41^ Numerous studies have demonstrated racial/ethnic and socioeconomic disparities in public drinking water exposures across the US, and have called for justice-oriented interventions and policies that protect the most highly exposed communities. While advancing environmental justice necessitates that all lead service lines should be identified and replaced within NYC and NYS, equitable efforts should prioritize the most impacted communities. Improvements to public health demands attention to all potential lead sources, including water, soil, and paint.

## Data Availability

All source data are available through the US Census Bureau and the New York City Open Data portal.

